# Accounting for Structured Missingness in Canonical Correlation Analysis

**DOI:** 10.1101/2025.10.09.25337581

**Authors:** Lav Radosavljević, Stephen M. Smith, Thomas E. Nichols

## Abstract

A particularly challenging form of missing data is structured missingness, where sets of subjects and variables consistently have missing data. For tabular data from sub-studies or modalities, structured missingness can come from non-participation in followup studies, which creates large blocks of missing data. Canonical Correlation Analysis (CCA) is a multivariate modelling tool commonly used to link two different set of variables, and in neuroimaging has typically been used to find associations between imaging and non-imaging variables. Motivated by CCA, we propose a new method for covariance estimation from incomplete data that handles data with a mix of structured and unstructured missingness, assuming Missing at Random (MAR). Our proposed method is compared to existing methodology by way of evaluation on simulated data and on real data from subjects in the UK Biobank brain imaging cohort.

## 1 Introduction

There has recently been an increased interest in studying and characterising the structure of missingness patterns in large-scale data sets^1–3^. Structured Missingness (SM) can have diverse causes, such as combining sub-studies with overlapping sets of participants, batch failure or recording data from a questionnaire where some questions are recorded only if the subject answered ‘Yes’ or ‘No’ to a previous question, leading to blocks of missing data^1^. While missing data literature mostly ignores missing data structure, this might be problematic. It is not uncommon for SM to lead to a situation where highly correlated variables are mostly jointly missing, making it much more difficult to impute missing values using correlated variables^3^. Therefore, new missing data methodology is needed to address SM specifically.

In this work, we focus on handling SM for the task of Canonical Correlation Analysis (CCA)^4^. Our work is inspired by tabular data from subjects in the UK Biobank (UKB) Brain Imaging Cohort. This data consists of imaging derived phenotypes (IDPs), which are derived from MRI images of the subjects’ brains, and non-imaging derived phenotypes (nIDPs), which come from diverse sub-studies in UKB. SM in this type of data is caused by non-participation, as each subject has participated in a specific set of sub-studies, and non-participation in any sub-study will lead to missingness for all variable derived from the same. Additionally, a sub-study can also have unstructured missingness (UM) that is attributable to any other cause. Figure 1 illustrates this setting, which is similar to a multi-view learning problem with missing views^5,6^, and with UM added. CCA is commonly used to link IDPs and nIDPs, both of which can have SM, so it is therefore important to investigate how to best handle SM for this analytical task. We evaluate several existing methods on both real and synthetic data and propose a new method which handles SM and UM separately.

**Figure 1.**
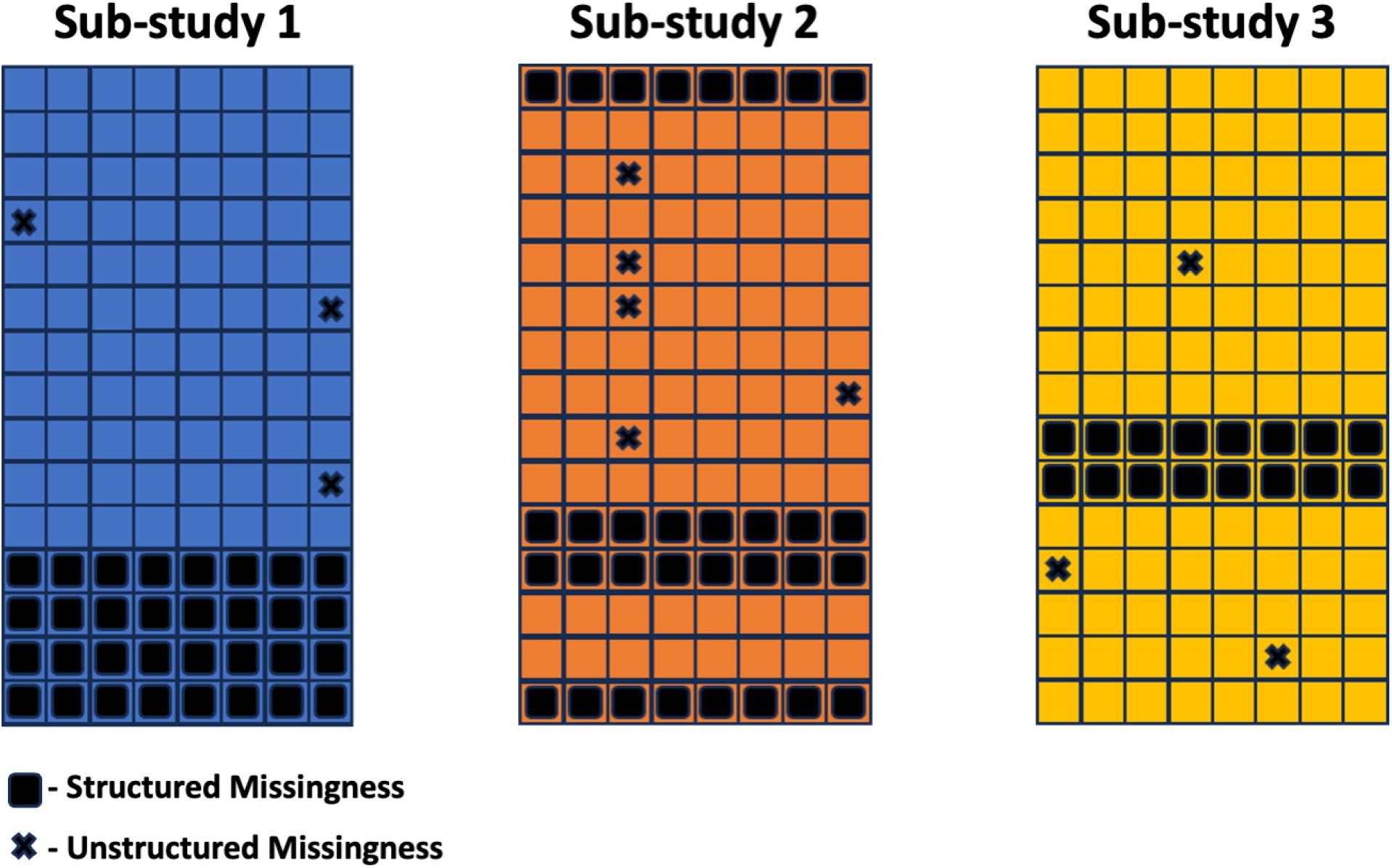
The missing data in a sub-study can be divided into structured missingness (SM), caused by non participation, and unstructured missingness (UM) attributable to any other cause.

### 1.1 Canonical Correlation Analysis

CCA is a multivariate analysis method commonly used in e.g. neuroscience for tasks such as de-noising, detecting task fMRI activation or analysing and testing associations between different modalities^7–11^. It finds the maximal correlation between linear combinations of two variable sets. If 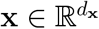 and 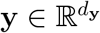are random vectors representing the two variable sets, CCA seeks to find weight vectors 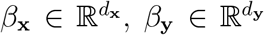such that *canonical variables u* = **x**^⊤^*β*_**x**_ and *v* = **y**^⊤^*β*_**y**_ have maximal correlation. CCA can be performed via the generalised eigenvalue problem (GEP) in the following way^12^:

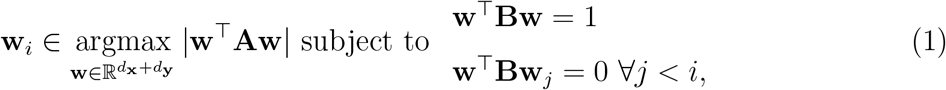

Where 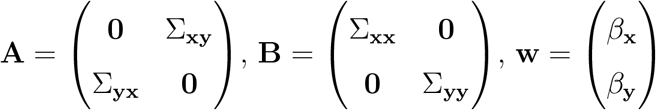and 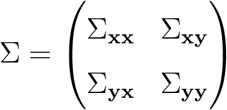 is the covariance matrix of (**x, y**).

In practice, since the joint covariance matrix of **x** and **y** is usually unknown, it is estimated from data; *D* = (**X, Y**), where **X** and **Y** are *n* × *d*_**x**_ and *n* × *d*_**y**_ data matrices corresponding to *n* independent observations of (**x, y**).

### 1.2 Confounding and Missingness

When analysing associations between IDP and nIDP data, there are often sources of confounding which we want to adjust for prior to performing CCA. In epidemiological studies, variables such as age, sex, BMI, ethnicity, geographical region and variables related to socioeconomic status are examples of confounds^13^. These are the same types of variables that are also broadly predictive of study participation^14^ and therefore also SM. Figure 2 shows the relationship between missingness and confounding variables that is assumed throughout the paper. Also, if we assume that our confound variable **b** is fully observed, we see that this missing data model falls under *Missing at Random* (MAR), since the relationship between data can be described using only observed variables. If **m** is the missingness indicator of our joint random vector **d** = (**b, x, y**), which can be split into observed and unobserved components **d**_obs_ and **d**_mis_, then we have that

**Figure 2:**
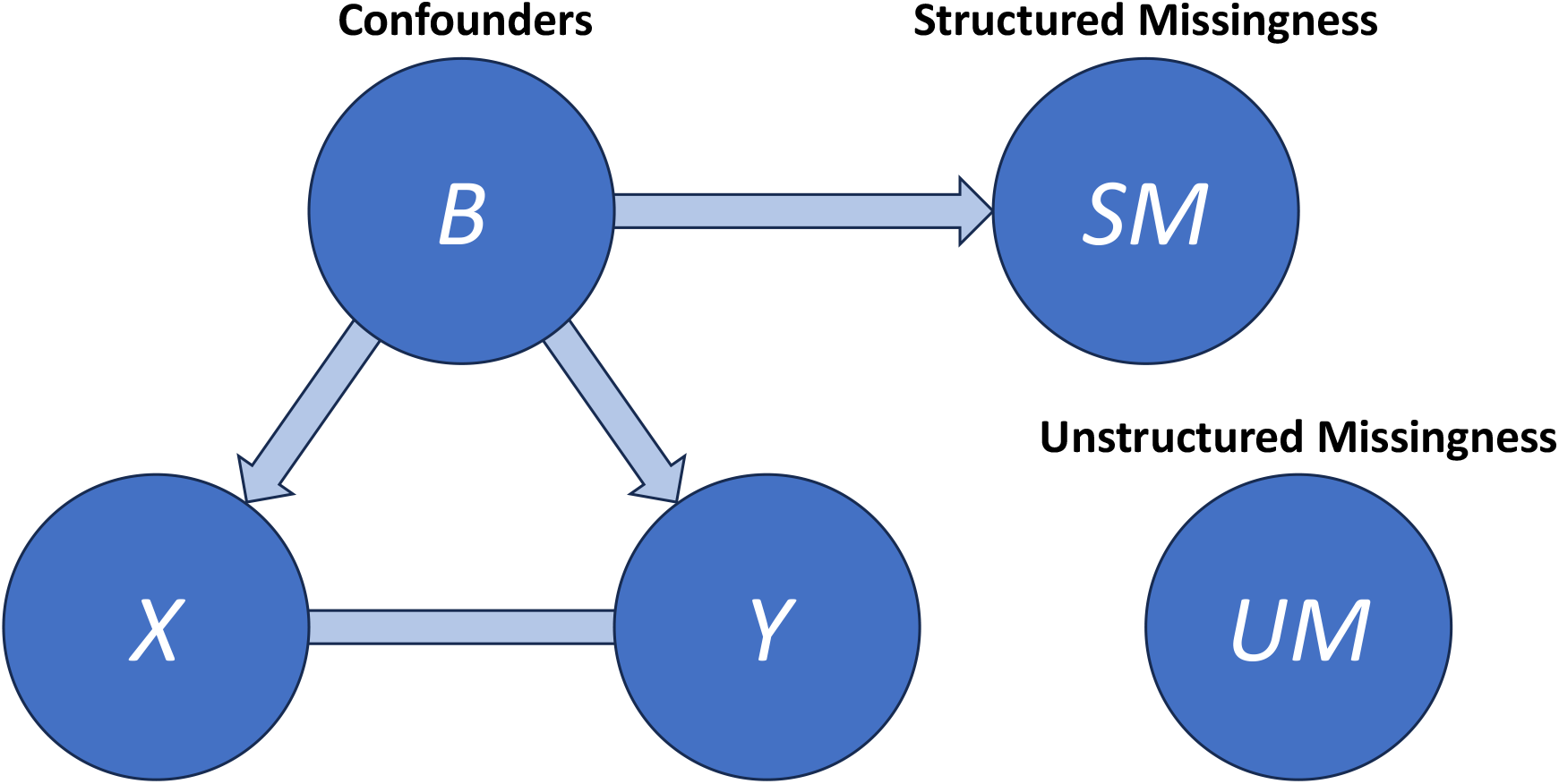
Structured missingness is assumed to be determined by the confounding variables, i.e., baseline characteristics such as age, sex, BMI, ethnicity, socioeconomic status etc., while unstructured missingness is assumed to be independent of all variables. This assumption results in data that falls under MAR, which our method relies on.

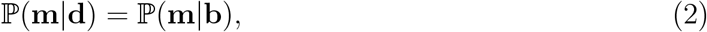

and since **b** *⊆* **d**_obs_ (confounds being always fully observed), it follows that

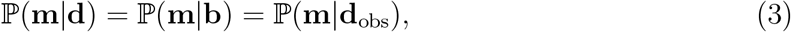

which by definition means that data is MAR.

### 1.3 Related Work

Since CCA requires estimation of the covariance matrix from observed data, the work most relevant to this paper concerns the estimation of covariance matrices from incomplete data. Under the assumption of multivariate Gaussianity and MAR, the Maximum Likelihood Estimate (MLE) of the covariance matrix Σ can be found via the Expectation-Maximisation (EM) algorithm^15^. This is, however, a highly computationally expensive procedure, as each iteration of the EM algorithm requires *h* inversions of (*d*_**x**_ + *d*_**y**_) × (*d*_**x**_ + *d*_**y**_) matrices, where *h* is the number of missingness patterns observed in the *n* rows of *D*. It is easy to see that any unstructured missingness will lead to *h* ≈ *n* in the high dimensional case, because there are 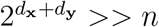 possible missingness patterns **m**, and it is therefore unlikely that the same missingness pattern would appear more than once. This consequently leads to a very high time complexity for the EM algorithm. Therefore, the use of the MLE of the covariance matrix via EM has mostly been restricted to low-dimensional, small data settings^16^.

Several faster unbiased methods using inverse probability weighting (IPW) have been proposed for learning covariance matrices from incomplete data assuming *missing completely at random* (MCAR). The method proposed by Lounici (2014)^17^ assumes that missingness for each entry is determined by independent and identically distributed Bernoulli random variables with a known probability *δ*. However, this estimator is heavily biased in our structured setup where there is both strong dependence between missing data patterns and differing rates of missingness between variables. Park and Lim (2019)^18^ extend this approach to allow for dependence between missingness patterns and variable-specific missingness rates, but their proposed estimator requires a-priori knowledge of the joint probability distribution of the missingness over all variables. If these parameters are estimated from data, the Park and Lim estimator becomes the standard covariance matrix estimate using pairwise complete observations (PCO), which is not necessarily positive semi-definite (PSD).

Multivariate Imputation by Chained Equations (MICE) ^19,20^ is also commonly used to handle missing data and is applicable to the problem of covariance estimation from incomplete data. MICE iteratively creates random synthetic replacements for the missing entries by fitting regression models for each variable using all other variables, which stochastically impute missing entries. Like the EM algorithm, this method also suffers from high computational cost, although this problem can be alleviated by using a carefully selected subset of variables to fit the prediction models, instead of using all available variables^3,21–23^.

Finally, it should be noted that in most cases, missing data is handled via simple, non-stochastic, Single Imputation methods such as mean imputation or matrix completion, prior to applying CCA. This is an inherently biased approach, unless imputation can be done with perfect accuracy, as non-stochastic imputation creates a downward bias in variance by not accounting for noise in the imputed values. And since the variance is biased post imputation, covariance is also generally biased.

## 2 Methods

Our method handles structured and unstructured missingness separately. When linking two modalities using CCA, we are often very close to a pure multi-view setting (i.e., a setting with no UM) where the EM algorithm can be practically used for covariance matrix estimation. However, the unstructured missingness makes this intractable, as discussed in subsection 1.3 and also in Le Morvan et. al. (2020)^24^. Additionally, we assume that correlations within **X** and **Y** are strong, while correlations between **X** and **Y** are weak, making unstructured missingness easy to impute and structured missingness difficult to impute. For example, a study measuring the weights of different body parts will produce highly correlated variables, as the weights of individual body parts are strongly correlated with the subjects’ height and BMI. Variables from different studies that measure completely different things tend to be more weakly correlated. We therefore propose to handle unstructured missingness prior to applying the EM algorithm, either via a single imputation method such as mean imputation or SoftImpute (SI)^25^, or via MICE. After that, we use EM to obtain an approximate MLE of the covariance matrix from incomplete data. In Figure 3, we see an illustration that describes the estimation procedure. When using MICE, we create *r* multiply imputed data sets, obtain MLE covariance matrices for each of them, and average them to obtain a single estimate.

**Figure 3:**
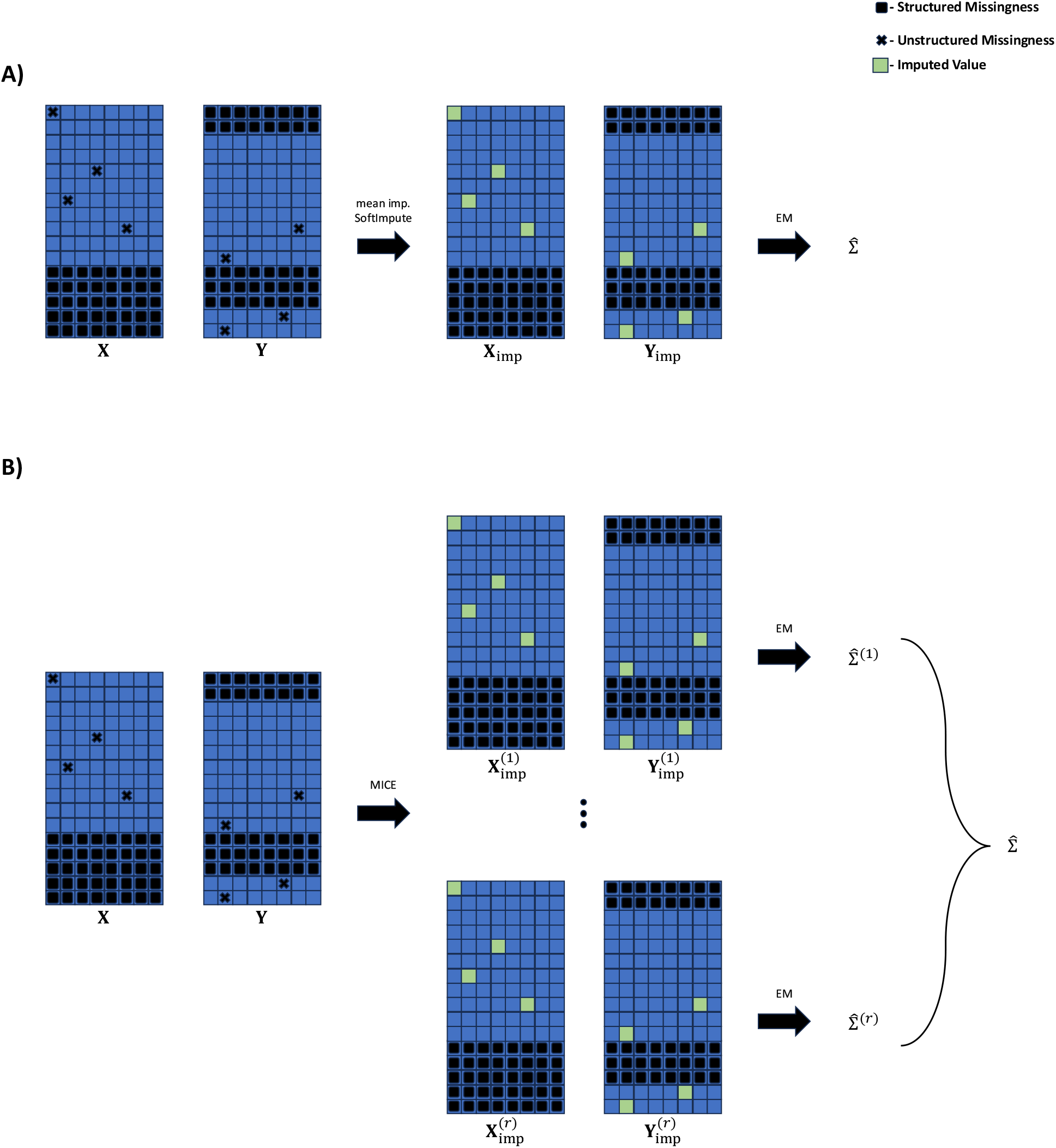
Illustration of our proposed method. A) We apply a Single Imputation method to impute the unstructured missingness in **X** and **Y**. After this, the EM algorithm is applied to obtain the MLE covariance matrix. B) We use MICE to create *r* multiply imputed data sets where only unstructured missingness has been imputed. We apply EM to obtain *r* MLE covariance matrices, which we average to obtain a single estimate.

There is mathematical intuition to support the validity of the proposed method using MICE. Let Σ be the covariance matrix described in equation 1. If *D*_obs_ = (**X**_obs_, **Y**_obs_) is the observed data and *D*_UM_ = (**X**_UM_, **Y**_UM_) is unobserved data obscured by unstructured missingness only, we know that

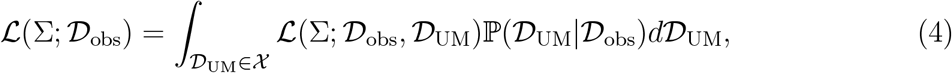

where ℒ is the likelihood function. It can be argued further, using the logic of Monte-Carlointegration that

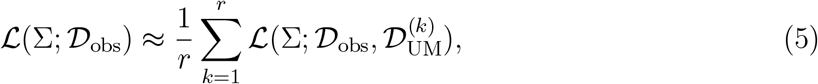

where 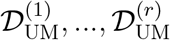 have been sampled independently from ℙ(*D*_UM_|*D*_obs_) using MICE. It follows that

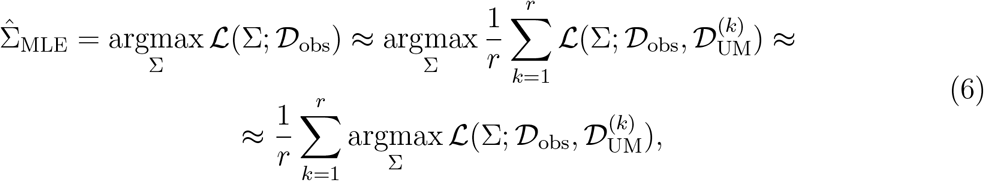

where the second approximation assumes that the functions 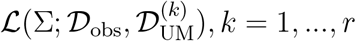are well approximated by quadratic functions with the same Hessian (see supplementary material for a full justification). Since *D*_obs_ and 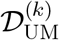 together form an imputed data set with only structured missingness, it is computationally tractable to find argmax_Σ_ 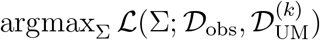 via EM. Thus, our final estimate is

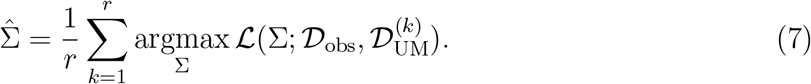

In practice, when finding the average of covariance matrices, they are often averaged in a reparametrised form and then returned to their original form after averaging^26,27^. We average the covariance matrices in Cholesky space^28^, because this is a simple and fast reparametrisation that has been shown to have good empirical performance for covariance matrices that are well conditioned^27^. This is a version of the standard method of pooling multiply imputed data, called Rubin’s rules^19^, performed on covariance matrices in a reparametrised form.

In order to save computational time, we only use the variables in **X** to impute **X**_UM_ and only the variables in **Y** to impute **Y**_UM_, as variables from the same sub-study tend to be more strongly correlated than variables from different substudies, making them more suitable for imputation. For the version of our method that uses MICE to impute UM, we only use a selection of 5 imputation variables for each variable.

The main benefit of our proposed method is that the covariance matrix is estimated efficiently without imputation, after handling the small amount of UM. The EM MLE after imputing UM uses all data available, in contrast with MICE where we are forced to select a subset for each variable that we use for imputation. This both saves computational time and reduces bias by avoiding this type of truncation.

### 2.1 Other Methods

We compare several other methods to our approach. The simplest of these include Single Imputation methods, mean imputation and SoftImpute. For SoftImpute, we select the soft threshold for singular values heuristically using the elbow rule^29^. Another method is MICE, where we create *r* multiply imputed data sets, calculate the sample covariance matrix on each set, and average them to obtain a single estimate (pooling is done in Cholesky space as described in section 2). Since we are working in a high dimensional setting, we use the variable selection procedure for iterative imputation models described in Radosavljević et. al. (2024)^3^. Here, we use 20 imputation variables for each variable. Finally, we consider the method of estimating the covariance matrix using pairwise complete observations. This estimate is unbiased under MCAR, but is not necessarily PSD. We solve this by first calculating standard deviations for each variable using complete observation. We then calculate the correlation matrix using the PCO approach and project it to the closest PSD correlation matrix with Higham’s Algorithm^30^ using the nearPD function in R’s Matrix package^31^. Finally, we transform this correlation matrix back to a PSD covariance matrix using the estimated standard deviations.

We acknowledge that there could be some benefit in excluding rows that have structured missingness in either **X** or **Y**, especially for Single Imputation methods where bias is increased with the amount of missingness. Assuming that the causal structure shown in Figure 2 is correct, excluding rows with SM should yield correct inferential results, as fully observed confounds contain all information that is predictive of missingness. On the other hand, the loss of information by excluding whole rows with SM could be detrimental to inference, or lead to a situation where the number of rows and columns are similar, which is itself problematic for weight stability in CCA. We therefore perform all methods (excluding our proposed method) using both approaches, as shown in Figure 4.

**Figure 4:**
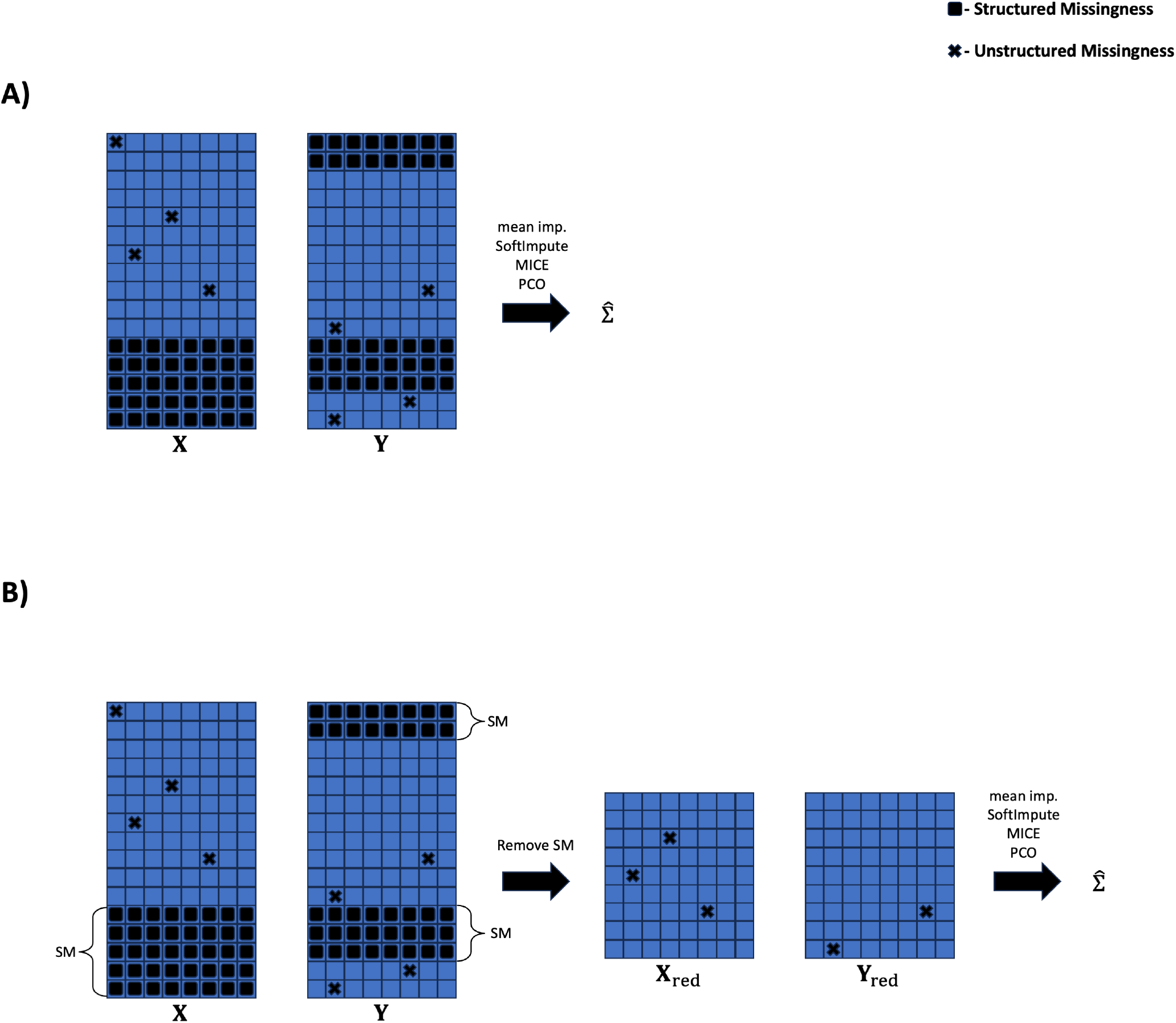
Illustration of the two approaches used for CCA. A) We use all rows in **X** and **Y**, irrespective of structured missingness. B) All rows where there is SM in either **X** or **Y** are excluded prior to downstream analysis.

### 2.2 Deconfounding

To adjust for confounding variables in our CCA, we first exclude the few rows where there is any missingness in the confounds and then perform standard Ordinary Least Squares (OLS) based deconfounding on **X** and **Y** as a preprocessing step. This is a standard approach to handling confounds in CCA^32^. Interestingly, since we assume that structured missingness is determined in large part by confounds, our data will be approximately MCAR after deconfounding, as the data in **X** and **Y** will be orthogonal to the confounds, and thus by extension uncorrelated with the structured missingness patterns.

### 2.3 Simulation Study

To evaluate our method, we perform a simulation study mimicking the real data case. We simulate data sets *D* = (**B, X, Y**) where **B** are confounds and (**X, Y**) is the pair of substudies that we want to perform CCA on. The process for generating confounds is shown using pseudo-code in the supplementary material. The data generation process is inspired by Singular Value Decomposition (SVD) and assumes the existence of a low rank approximation of data. (**X, Y**) are generated in a similar way, where 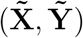 are two matrices with common variance generated using an SVD inspired procedure, and we set

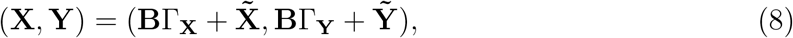

where Γ_**X**_ and Γ_**Y**_ are two matrices of confound effects.

We induce structured missingness in (**X, Y**) using our simulated **B**, such that the probability of SM in **X** is *p*_SM_ and 1.5*p*_SM_ in **Y**, while guaranteeing that SM can be predicted using **B** with an AUC score of AUC_SM_ = 0.8. The process we use for creating informative SM is described fully in Radosavljević et. al. (2024)^3^. SM is determined via a Logistic Regression model using the confounds as variables. The coefficients of the Logistic Regression model are scaled using a binary search procedure to match the AUC score that we have chosen (AUC_SM_). We also induce unstructured missingness at a rate of *p*_UM_ in total, but such that the probability of UM is allowed to vary between variables. Specifically, the rates of UM for each variable are drawn from a Beta distribution with expectation *p*_UM_ and variance 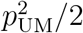Lastly, we induce 1% missingness in the confounds for a subset of variables that are not used to induce structured missingness.

We vary the rates of missingness to be *p*_SM_ = 0.1, 0.2, 0.4 and *p*_UM_ = 0.01, 0.05, 0.15. We also vary a parameter *σ*_*U*_ = 0.02, 0.03, 0.05, which controls the level of correlation between **X** and **Y** after removing confounding effects, with higher values of *σ*_*U*_ leading to stronger canonical correlation. *σ*_*U*_ is the standard deviation of the variance components that are shared between 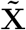and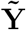, while the variance components that are unique to either 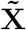 or 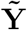 all have standard deviation 1.

We set *n* = 50000 (roughly the number of subjects in the UKB Brain Imaging cohort) and *d*_**x**_ = *d*_**y**_ = 200.

We evaluate the performance of each missingness method on 100 synthetic data sets for each setting, by estimating canonical variable coefficients on a training set and then calculating the average correlation of the first ten canonical variables constructed on a non-confounded test set using our estimated coefficients. In other words, if 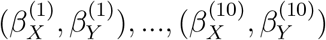 are our estimated canonical coefficients and 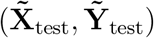 is our non-confounded test set, the score is:

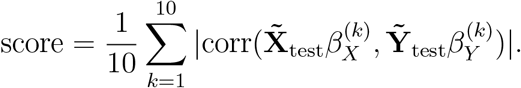

The goal is for this score to be as high as possible. We then obtain relative scores by dividing by the score obtained using the Gold Standard (GS), which in our case means analysis on data with no missingness. The relative scores (score*/*score_GS_) are thus usually in the interval (0, 1) and represent how close our missingness method gets us to replicating a scenario with no missingness. Given the assumptions of our method, we expect it to perform the best compared to other methods for cases where the rate of SM is high and the rate of UM is low, as UM has the lowest impact on covariance estimation in that scenario.

### 2.4 Study on Real Data

To evaluate our method on real data, we analyse four sub-studies from UK Biobank which have structured missingness. They include diffusion MRI (dMRI) weighted-mean tract diffusivity IDPs, food preferences, NMR metabolomics, and refractometry. We also include a set of confounds: sex, age, ethnic background (binary White British vs. not White British), income, townsend deprivation index, BMI, frequency of alcohol consumption, smoking status and geographical north and east coordinates. Additionally, we include age-squared, ageby-sex and age-squared-by-sex, to account for non-linear age effects and interaction effects between sex and age. Variable numbers, rates of SM and UM, as well as informativeness of SM for the data sets are shown in table 1. A table showing the proportion of overlapping participants for each pair of studies is shown in table 2. The sets are chosen to represent typical analyses of this kind: The IDP data is high-dimensional and has low rates of SM and UM, the food preferences data has moderate rates of SM and almost no UM, NMR metabolomics has a high rate of SM and low rate of UM, and the Refractometry variables have high levels of SM and UM. Table 2 also shows the proportion of subjects that have participated in both studies for each pair of studies, showing diverse settings ranging from 77% for IDPs vs. Food Preferences, to 5.5% for NMR metabolomics vs. Refractometry.

**Table 1:** Number of variables, level of structured (*p*_SM_) and unstructured (*p*_UM_) missingness as well as informativeness of structured missingness (AUC_SM_). AUC_SM_ is estimated by fitting a logistic regression model for predicting SM using confounds; AUC_SM_ is the AUC score for prediction calculated on test data.

| Set | #Variables | $p_{\text{SM}}$ | $p_{\text{UM}}$ | $\text{AUC}_{\text{SM}}$ |
| --- | --- | --- | --- | --- |
| Confounds | 21 | 0 | 0.002 | NA |
| IDPs | 300 | 0.053 | 0.012 | 0.61 |
| Food Preferences | 130 | 0.19 | $2.4 \cdot 10^{-6}$ | 0.57 |
| NMR metabolomics | 73 | 0.76 | 0.0029 | 0.51 |
| Refractometry | 96 | 0.77 | 0.095 | 0.74 |

**Table 2:**
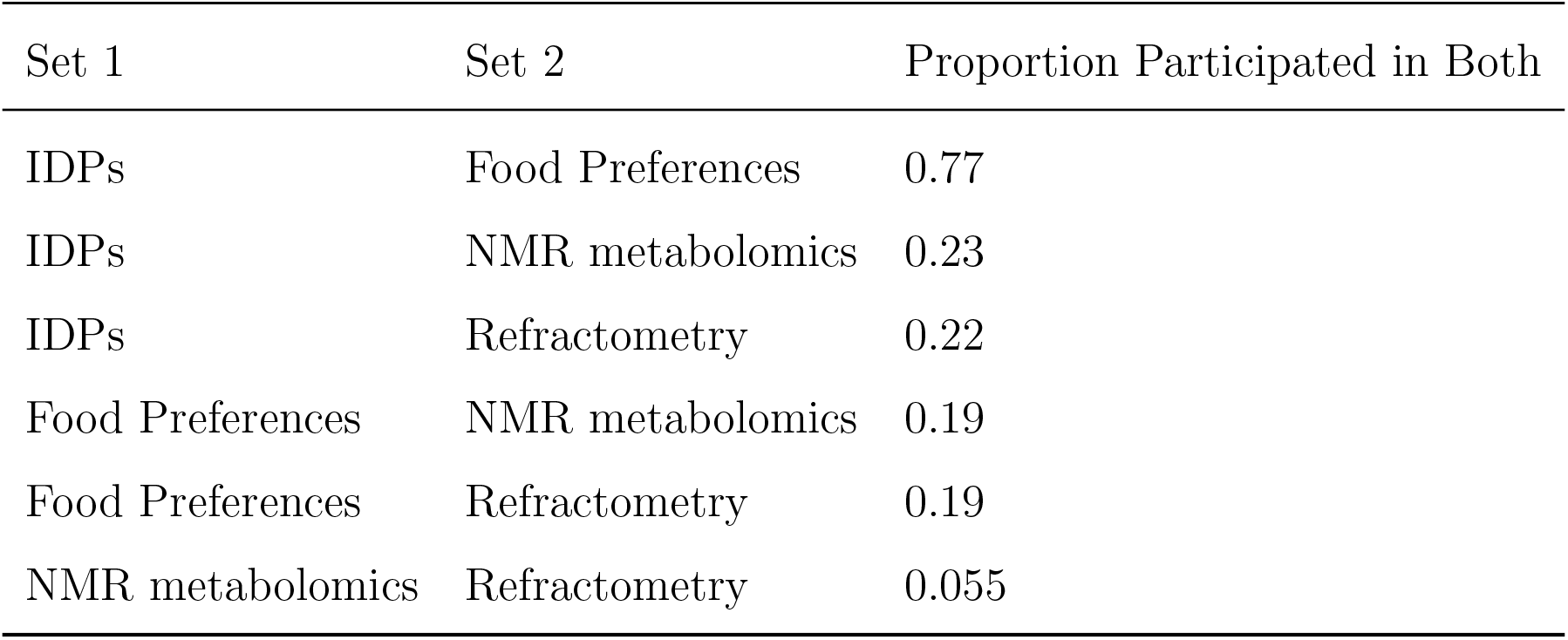
Proportion of subjects (out of *n* = 46471), that participated in both studies. The table shows a wide range of settings for CCA from structured missing data; for IDPs vs. Food Preferences, a comfortable majority of subjects participated in both studies, while for NMR metabolomics vs. Refractometry, fewer than 6% of subjects participated in both studies, and is consequently a much more unstable setting.

| Set 1 | Set 2 | Proportion Participated in Both |
| --- | --- | --- |
| IDPs | Food Preferences | 0.77 |
| IDPs | NMR metabolomics | 0.23 |
| IDPs | Refractometry | 0.22 |
| Food Preferences | NMR metabolomics | 0.19 |
| Food Preferences | Refractometry | 0.19 |
| NMR metabolomics | Refractometry | 0.055 |

We evaluate missing data methods for CCA on each of six possible pairs of sub-studies. For each pair of sub-studies, we make 100 train-test splits (80% *−* 20%). Deconfounded canonical coefficients are estimated on training data using each missingness method. On test data, the complete cases are kept, deconfounded and the canonical variables are calculated using CCA coefficients from each method. The correlation between these canonical variables constructed on test data is calculated and, with best methods having largest correlation.

We also show a real world example of how our method is be used to link two studies, food-preferences and NMR metabolomics, by applying our proposed method using EM combined with SoftImpute to find the first canonical modes of the two datasets. The canonical modes are then correlated with the variables in each respective study to find links between variables from different studies.

## 3 Results

### 3.1 Simulated Data

Table 3 shows the results of the simulation study described in subsection 2.3, detailing the performance of all methods relative to the Gold Standard under different simulation settings. We see that for most settings, the two versions of our proposed method that use SoftImpute and MICE (MICE+EM and SI+EM), have results comparable to those of MICE or better. This is particularly encouraging for the version that uses SoftImpute, as this is a very computationally cheap method. Our proposed methods give very similar performance to MICE’s and dominate in a few settings. The PCO approach performs by far the worst overall. Figure 5 shows the computational time of each method on a single synthetic data set. Our method is much faster than MICE, especially when using mean imputation and SoftImpute to handle UM. MICE takes ≈ 2 hours in total, whereas MICE+EM takes ≈ 40 minutes, and mean+EM and SoftImpute+EM both take below 1 minute.

**Table 3:** Simulation results showing the performance of each method in different settings. The results in each entry show the average of the first 10 canonical correlations on held out data relative to the same average for the Gold Standard method, which is the case where we have no missing data (see subsection 2.3). The median result over 100 simulations is recorded. Bold font indicates the best result for a given setting. We see that the versions of our method that use SoftImpute or MICE to impute UM perform competitively compared to MICE.

| Setting |  |  | SM Excluded |  |  |  | SM Not Excluded |  |  |  |  |  |  |
| --- | --- | --- | --- | --- | --- | --- | --- | --- | --- | --- | --- | --- | --- |
| $p_{UM}$ | $p_{SM}$ | $\sigma_U$ | mean | PCO | SI | MICE | mean | PCO | SI | MICE | mean+EM | SI+EM | MICE+EM |
| 0.01 | 0.1 | 0.05 | 0.835 | 0.832 | 0.839 | 0.838 | 0.834 | 0.832 | 0.836 | **0.844** | 0.834 | 0.839 | 0.838 |
|  |  | 0.03 | 0.678 | 0.672 | 0.683 | 0.687 | 0.679 | 0.670 | 0.676 | **0.698** | 0.673 | 0.686 | 0.677 |
|  |  | 0.02 | 0.569 | 0.545 | 0.574 | 0.574 | 0.565 | 0.541 | 0.543 | **0.596** | 0.571 | 0.589 | 0.573 |
|  | 0.2 | 0.05 | 0.807 | 0.804 | 0.809 | 0.809 | 0.805 | 0.800 | 0.801 | **0.811** | 0.807 | 0.809 | 0.809 |
|  |  | 0.03 | 0.601 | 0.583 | 0.614 | 0.617 | 0.600 | 0.577 | 0.570 | **0.636** | 0.600 | 0.615 | 0.614 |
|  |  | 0.02 | 0.490 | 0.466 | 0.507 | 0.508 | 0.496 | 0.471 | 0.467 | **0.519** | 0.483 | 0.506 | 0.500 |
|  | 0.4 | 0.05 | 0.737 | 0.725 | 0.744 | **0.745** | 0.730 | 0.717 | 0.652 | 0.709 | 0.735 | 0.742 | 0.742 |
|  |  | 0.03 | 0.435 | 0.400 | 0.432 | 0.443 | 0.423 | 0.406 | 0.284 | 0.437 | 0.410 | 0.439 | **0.445** |
|  |  | 0.02 | 0.323 | 0.292 | 0.321 | **0.337** | 0.311 | 0.315 | 0.205 | 0.302 | 0.308 | 0.330 | 0.326 |
| 0.05 | 0.1 | 0.05 | 0.803 | 0.766 | 0.824 | 0.826 | 0.802 | 0.766 | 0.820 | 0.830 | 0.803 | 0.825 | 0.820 |
|  |  | 0.03 | 0.639 | 0.496 | 0.670 | 0.679 | 0.643 | 0.495 | 0.665 | **0.678** | 0.639 | 0.672 | 0.675 |
|  |  | 0.02 | 0.502 | 0.344 | 0.533 | **0.554** | 0.494 | 0.355 | 0.510 | 0.547 | 0.498 | 0.544 | 0.547 |
|  | 0.2 | 0.05 | 0.793 | 0.731 | 0.808 | 0.810 | 0.791 | 0.729 | 0.802 | 0.806 | 0.792 | **0.811** | 0.808 |
|  |  | 0.03 | 0.576 | 0.432 | 0.613 | 0.621 | 0.575 | 0.436 | 0.598 | **0.631** | 0.577 | 0.615 | 0.613 |
|  |  | 0.02 | 0.455 | 0.320 | 0.481 | 0.487 | 0.445 | 0.320 | 0.431 | **0.496** | 0.442 | 0.488 | 0.482 |
|  | 0.4 | 0.05 | 0.659 | 0.493 | 0.681 | **0.687** | 0.648 | 0.504 | 0.562 | 0.672 | 0.659 | 0.682 | 0.681 |
|  |  | 0.03 | 0.358 | 0.222 | 0.377 | 0.389 | 0.356 | 0.236 | 0.235 | **0.423** | 0.349 | 0.372 | 0.377 |
|  |  | 0.02 | 0.301 | 0.217 | 0.332 | 0.342 | 0.298 | 0.236 | 0.217 | **0.348** | 0.309 | 0.335 | 0.338 |
| 0.15 | 0.1 | 0.05 | 0.777 | 0.494 | 0.811 | 0.811 | 0.776 | 0.499 | 0.809 | 0.810 | 0.776 | **0.817** | 0.800 |
|  |  | 0.03 | 0.549 | 0.203 | 0.620 | **0.629** | 0.549 | 0.210 | 0.608 | 0.629 | 0.549 | 0.627 | 0.608 |
|  |  | 0.02 | 0.419 | 0.205 | 0.478 | **0.516** | 0.419 | 0.209 | 0.453 | 0.501 | 0.427 | 0.487 | 0.502 |
|  | 0.2 | 0.05 | 0.714 | 0.326 | 0.753 | 0.765 | 0.709 | 0.342 | 0.741 | 0.757 | 0.714 | 0.762 | 0.752 |
|  |  | 0.03 | 0.467 | 0.157 | 0.528 | **0.545** | 0.484 | 0.162 | 0.469 | 0.533 | 0.467 | 0.540 | 0.518 |
|  |  | 0.02 | 0.379 | 0.164 | 0.418 | **0.453** | 0.378 | 0.167 | 0.378 | 0.430 | 0.376 | 0.442 | 0.418 |
|  | 0.4 | 0.05 | 0.595 | 0.129 | 0.644 | **0.669** | 0.588 | 0.166 | 0.492 | 0.634 | 0.594 | 0.655 | 0.657 |
|  |  | 0.03 | 0.283 | 0.079 | 0.304 | **0.351** | 0.284 | 0.091 | 0.187 | 0.364 | 0.274 | 0.320 | 0.337 |
|  |  | 0.02 | 0.251 | 0.132 | 0.269 | **0.295** | 0.240 | 0.134 | 0.206 | 0.284 | 0.237 | 0.261 | 0.272 |

**Figure 5:**
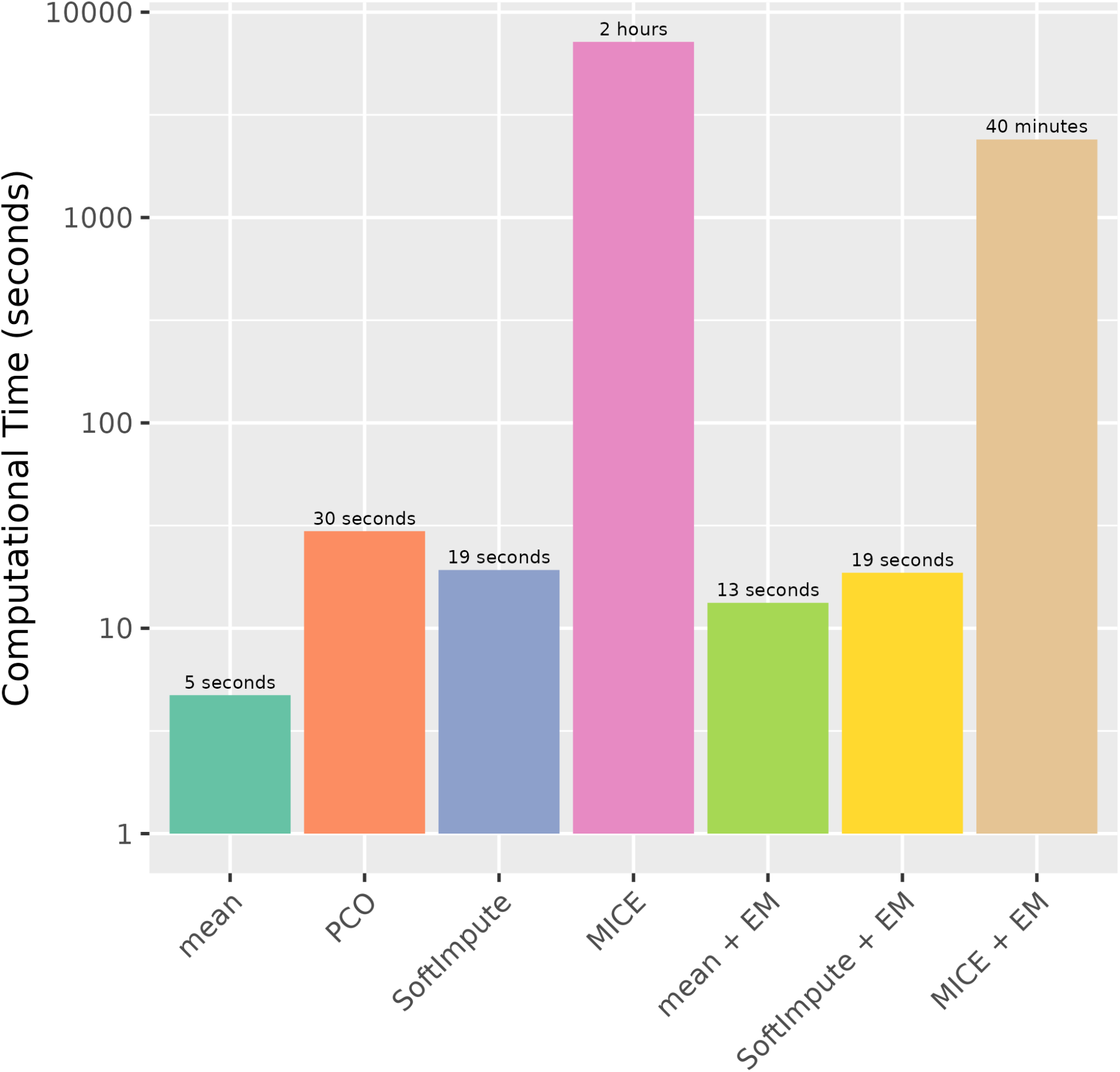
Computational time for each method on a single synthetic data set (*p*_SM_ = 0.2, *p*_UM_ = 0.05, *σ*_*U*_ = 0.03). The *y*-axis is shown on a logarithmic scale. We can see that our method is much faster than MICE, especially when using mean imputation and SoftImpute to impute UM.

### 3.2 Real Data

Table 4 shows the results on real data. As we can see, the three versions of our proposed method perform the best relative to other methods for pairs of data sets that include the IDPs. A reasonable explanation for this is that the IDPs have by far the most number of variables, and our proposed method is designed for high dimensional settings. Conversely, all three versions of our method have low scores for the NMR metabolomics and Refractometry pair. This can possibly be explained by the low level of correlation between the data sets, and low percentage of participants of both studies, leading to very unstable coefficients. For the two remaining comparisons, our methods perform similar to MICE. A possible explanation for this is that all other sets have ≈ 100 variables, so the dimensionality is sufficiently low for MICE to work almost optimally (see subsection 2.1 for details on “truncating” MICE in the high-dimensional setting). As with simulated data, we also see that the PCO approach performs the worst.

**Table 4:** Results for evaluation on real data. Bold font indicates the best result for a given pair of sub-studies. As we can see, MICE combined with EM has the best performance for all pairs of studies that involve the IDPs. A reasonable explanation for this is that the IDPs have by far the most number of variables, and our proposed method is designed for high dimensional settings. All three versions of our method have low scores for the NMR metabolomics and Refractometry pair. This can possibly be explained by the low level of correlation between the data sets, and low percentage of participants of both studies, leading to very unstable coefficients.

| Sub-studies |  | SM Excluded |  |  |  | SM Not Excluded |  |  |  |  |  |  |
| --- | --- | --- | --- | --- | --- | --- | --- | --- | --- | --- | --- | --- |
| Set 1 | Set 2 | mean | PCO | SI | MICE | mean | PCO | SI | MICE | mean+EM | SI+EM | MICE+EM |
| IDPs | Food Pref. | 0.215 | -0.003 | 0.217 | 0.217 | 0.214 | 0.000 | 0.217 | 0.217 | 0.215 | 0.216 | **0.218** |
| IDPs | NMR metabol. | 0.113 | 0.012 | 0.113 | **0.116** | 0.115 | 0.004 | 0.068 | 0.057 | 0.103 | 0.104 | **0.116** |
| IDPs | Refrac. | 0.068 | 0.013 | 0.077 | 0.100 | 0.054 | 0.003 | 0.036 | 0.065 | 0.062 | 0.070 | **0.113** |
| Food Pref. | NMR metabol. | 0.303 | 0.300 | 0.303 | 0.303 | 0.298 | 0.293 | **0.311** | 0.301 | 0.304 | 0.304 | 0.304 |
| Food Pref. | Refrac. | 0.012 | 0.000 | 0.014 | 0.025 | 0.011 | -0.007 | **0.044** | 0.033 | 0.004 | 0.000 | 0.015 |
| NMR metabol. | Refrac. | 0.215 | -0.013 | 0.214 | 0.230 | **0.240** | 0.041 | 0.056 | -0.001 | 0.111 | 0.120 | 0.177 |

In Figures 6 and 7, we see the variables most strongly correlated or anti-correlated with the first canonical modes of the food preference and NMR metabolomics datasets. The first canonical correlation between the two datasets is *r* = 0.38. As we can see, the main link between the two datasets seems to be the positive correlation between a preference for fatty fish, and Omega-3 and Docosahexaenoic fatty acids. The correlation between consuming fatty fish and Omega-3 is a well established results in nutrition science^33,34^. The equivalent plots for the five remaining CCA comparisons are found in the supplementary material.

**Figure 6:**
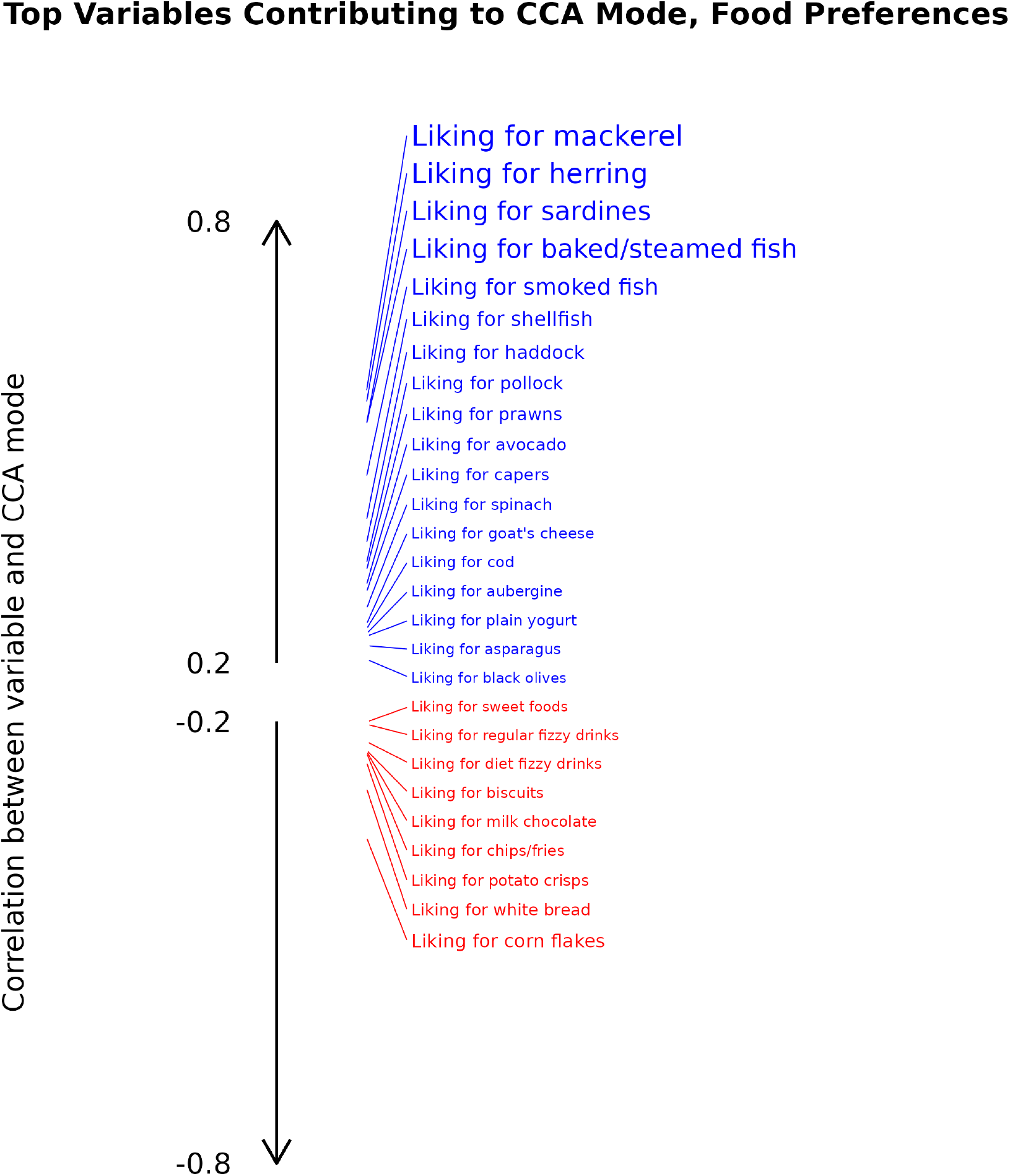
Food preferences: top variables in terms of association with the first canonical mode. We see a clear pattern; the canonical mode is positively correlated with a liking for fatty fish and other seafood, and negatively associated with a liking for a number of carbohydrate rich foods. The first canonical correlation is *r* = 0.38.

**Figure 7:**
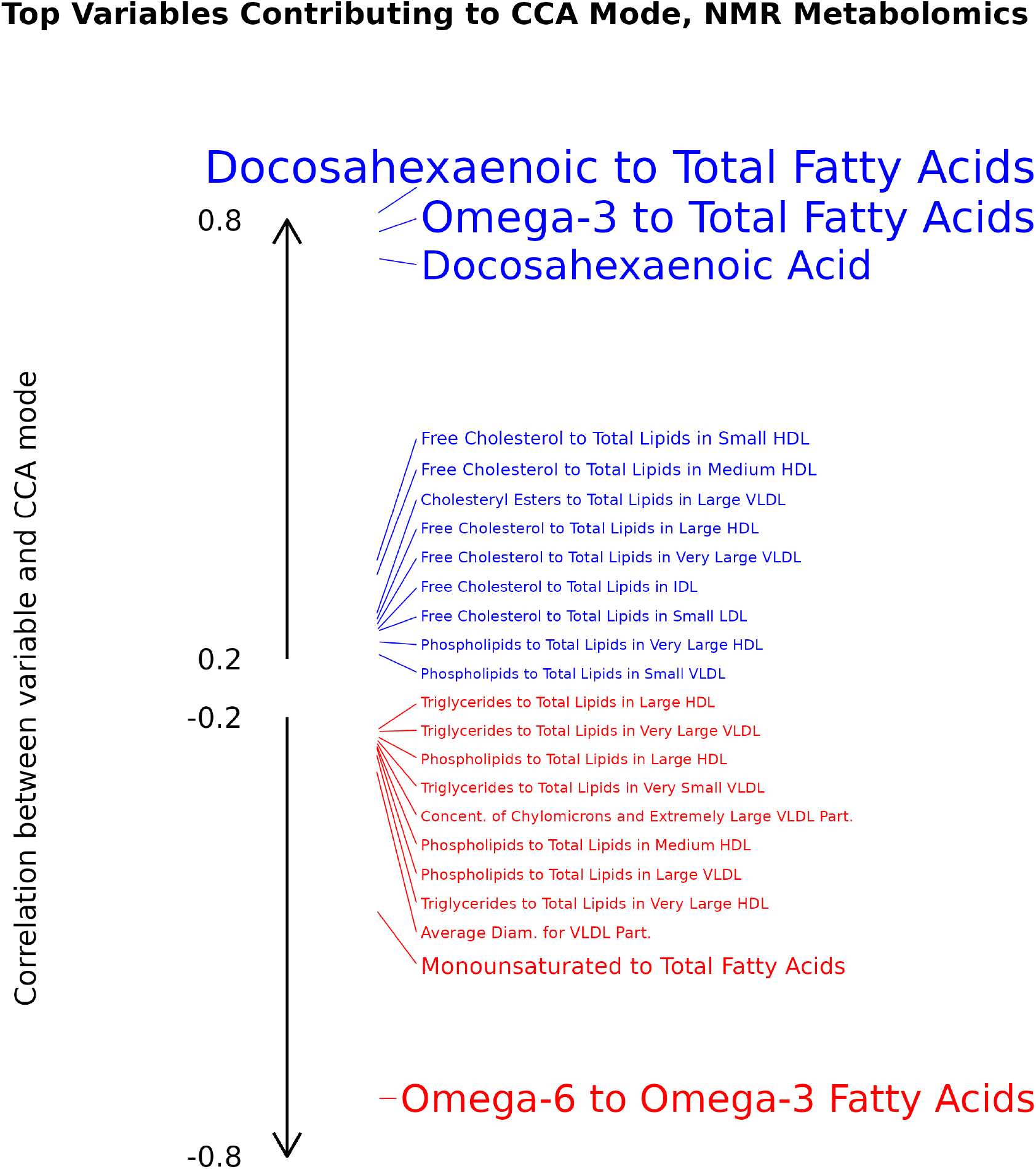
NMR metabolomics: top variables in terms of association with the first canonical mode. We see a very clear pattern; the canonical mode is strongly positively correlated with the proportion of Omega-3 and Docosahexaenonic fatty acids out of total fatty acids. The first canonical correlation is *r* = 0.38.

## 4 Discussion

We have proposed a new method for handling missing data for CCA. As seen from both real and simulated data, our proposed method is most useful relative to other methods in cases where the rate of UM is low, and we have also shown that it is computationally much faster than the best method, MICE. Conversely, we have shown that the proposed method performs poorly when there is a high level of UM. For those cases, SoftImpute or MICE on data where SM is excluded are better alternatives. For example, in a situation where we have two data sets with > 300 variables each, and the rate of UM is *<* 3% in both sets, while the rate of SM is > 20% in either of the sets, we strongly recommend the use of our method as compared to a more standard approach such as MICE, because it is much faster than MICE and at least as accurate.

In this work, we have limited ourselves to cases where CCA is performed on pairs of sub-studies. It is however, completely viable to use our proposed method for cases where we are dealing with multiple sub-studies, either as parts of **X** and **Y** or in the case of multi-set CCA (MCCA)^35,36^, i.e., an extension of CCA with multiple data sets instead of merely two. In fact, it is entirely possible that our proposed method is more useful in these settings, as it becomes, in these cases, impossible to remove all rows with SM without reducing the number of rows to be either zero or lower than the total number of variables.

Lastly, we would like to note that our work shows that incorporating knowledge of SM in missing data methodology can yield better and faster results than standard approaches that are agnostic to structure. It is therefore necessary to study the effect of SM on analysis and develop suitable methodology for SM more broadly.

Our proposed method of covariance estimation and CCA has been released as an R package and can be found in the following GitHub repository: https://github.com/lavrad99/SM_CCA

## Data Availability

Simulated data can be made available upon reasonable request to authors.

## Declarations

### Ethics approval and consent to participate

The UK Biobank has received ethical approval from the North West Multi-Center Research Ethics Committee (11/NW/0382).

This research project received approval from the UKB under application number 8107.

This research project has adhered to the Declaration of Helsinki

## Acknowledgements

The computational aspects of this research were supported by the Wellcome Trust Core Award Grant Number 203141/Z/16/Z and the NIHR Oxford BRC. The views expressed are those of the author(s) and not necessarily those of the NHS, the NIHR or the Department of Health

