## Supplementary Material for "Accounting for Structured Missingness in Canonical Correlation Analysis"

#### Justification of Approximation

Assume that $f_i(\mathbf{x}) = \frac{1}{2}\mathbf{x}^\top \mathbf{A}\mathbf{x} + \mathbf{b}_i^\top \mathbf{x} + \mathbf{c}_i$, $f_i : \mathbb{R}^d \to \mathbb{R}$ for $i = 1, ...n$, is a series of quadratic functions with the same Hessian $\mathbf{A}$. Assume further that $A$ is negative definite, so that $f_i$ all have global maxima. We want to show that

$$\operatorname*{argmax}_{\mathbf{x}\in\mathbb{R}^d} \frac{1}{n}\sum_{i=1}^{n} f_i(\mathbf{x}) = \frac{1}{n}\sum_{i=1}^{n} \operatorname*{argmax}_{\mathbf{x}\in\mathbb{R}^d} f_i(\mathbf{x}).$$

It is easy to derive that

$$\operatorname*{argmax}_{\mathbf{x}\in\mathbb{R}^d} f_i(\mathbf{x}) = -\mathbf{A}^{-1}\mathbf{b}_i,$$

by setting the gradient of $f_i$ to $\mathbf{0}$. It therefore follows that

$$\frac{1}{n}\sum_{i=1}^{n} \operatorname*{argmax}_{\mathbf{x}\in\mathbb{R}^d} f_i(\mathbf{x}) = \frac{1}{n}\sum_{i=1}^{n} -\mathbf{A}^{-1}\mathbf{b}_i = -\mathbf{A}^{-1}\left(\frac{1}{n}\sum_{i=1}^{n}\mathbf{b}_i\right).$$

Let

$$\tilde{f}(\mathbf{x}) = \frac{1}{n}\sum_{i=1}^{n} f_i(\mathbf{x}) = \frac{1}{2}\mathbf{x}^\top\mathbf{A}\mathbf{x} + \left(\frac{1}{n}\sum_{i=1}^{n}\mathbf{b}_i\right)^\top \mathbf{x} + \left(\frac{1}{n}\sum_{i=1}^{n}\mathbf{c}_i\right).$$

$\tilde{f}$ is therefore also a quadratic function with Hessian $\mathbf{A}$, and it follows that

$$\operatorname*{argmax}_{\mathbf{x}\in\mathbb{R}^d} \tilde{f}(\mathbf{x}) = \operatorname*{argmax}_{\mathbf{x}\in\mathbb{R}^d} \frac{1}{n}\sum_{i=1}^{n} f_i(\mathbf{x}) = -\mathbf{A}^{-1}\left(\frac{1}{n}\sum_{i=1}^{n} \mathbf{b}_i\right).$$

$$\therefore \operatorname*{argmax}_{\mathbf{x}\in\mathbb{R}^d} \frac{1}{n}\sum_{i=1}^{n} f_i(\mathbf{x}) = \frac{1}{n}\sum_{i=1}^{n} \operatorname*{argmax}_{\mathbf{x}\in\mathbb{R}^d} f_i(\mathbf{x}).$$

Q.E.D

This lemma shows that our pooling of covariance matrices after MICE approximately corresponds to finding the MLE, if the likelihood functions for the covariance matrix of the multiply imputed datasets with only SM are well approximated by quadratic functions that have the same Hessian.

### Simulating Data

**Algorithm 1** Generating Confounds $\mathbf{B}$

$r \leftarrow \text{round}(0.3 d_B)$
**for** $i \leftarrow 1$ **to** $r$ **do**
  $s_i \sim \chi^2(1)$
**end for**
$s_{\max} \leftarrow \max\{s_1, s_2, \ldots, s_r\}$
**for** $i \leftarrow 1$ **to** $r$ **do**
  $s_i \leftarrow 0.05 + s_i / s_{\max}$
**end for**
**for** $i \leftarrow 1$ **to** $n$ **do**
  **for** $j \leftarrow 1$ **to** $r$ **do**
    $U_{ij} \sim \mathcal{N}(0, 1)$
  **end for**
**end for**
**for** $i \leftarrow 1$ **to** $d$ **do**
  **for** $j \leftarrow 1$ **to** $r$ **do**
    $V_{ij} \sim \mathcal{N}(0, 1)$
    $V_{ij} \leftarrow V_{ij}^3$
  **end for**
**end for**
$\mathbf{B} \leftarrow \mathbf{U}\text{diag}\{s_1, s_2, \ldots, s_r\}\mathbf{V}^\top$
$\Sigma_{\text{NONOISE}} \leftarrow \mathbf{V}\text{diag}\{s_1^2, s_2^2, \ldots, s_r^2\}\mathbf{V}^\top$
**for** $i \leftarrow 1$ **to** $n$ **do**
  **for** $j \leftarrow 1$ **to** $d$ **do**
    $t \sim \mathcal{N}\left(0, 1.5\{\Sigma_{\text{NONOISE}}\}_{jj}\right)$
    $B_{ij} \leftarrow B_{ij} + t$
  **end for**
**end for**

### CCA: Remaining Five Analyses

The plots for the remaining five pairs of CCA analyses are presented below in the following order:

1. IDPs vs. Food Preferences
2. IDPs vs. NMR Metabolomics
3. IDPs vs. Refractometry
4. Food Preferences vs Refractometry
5. NMR Metabolomics vs Refractometry

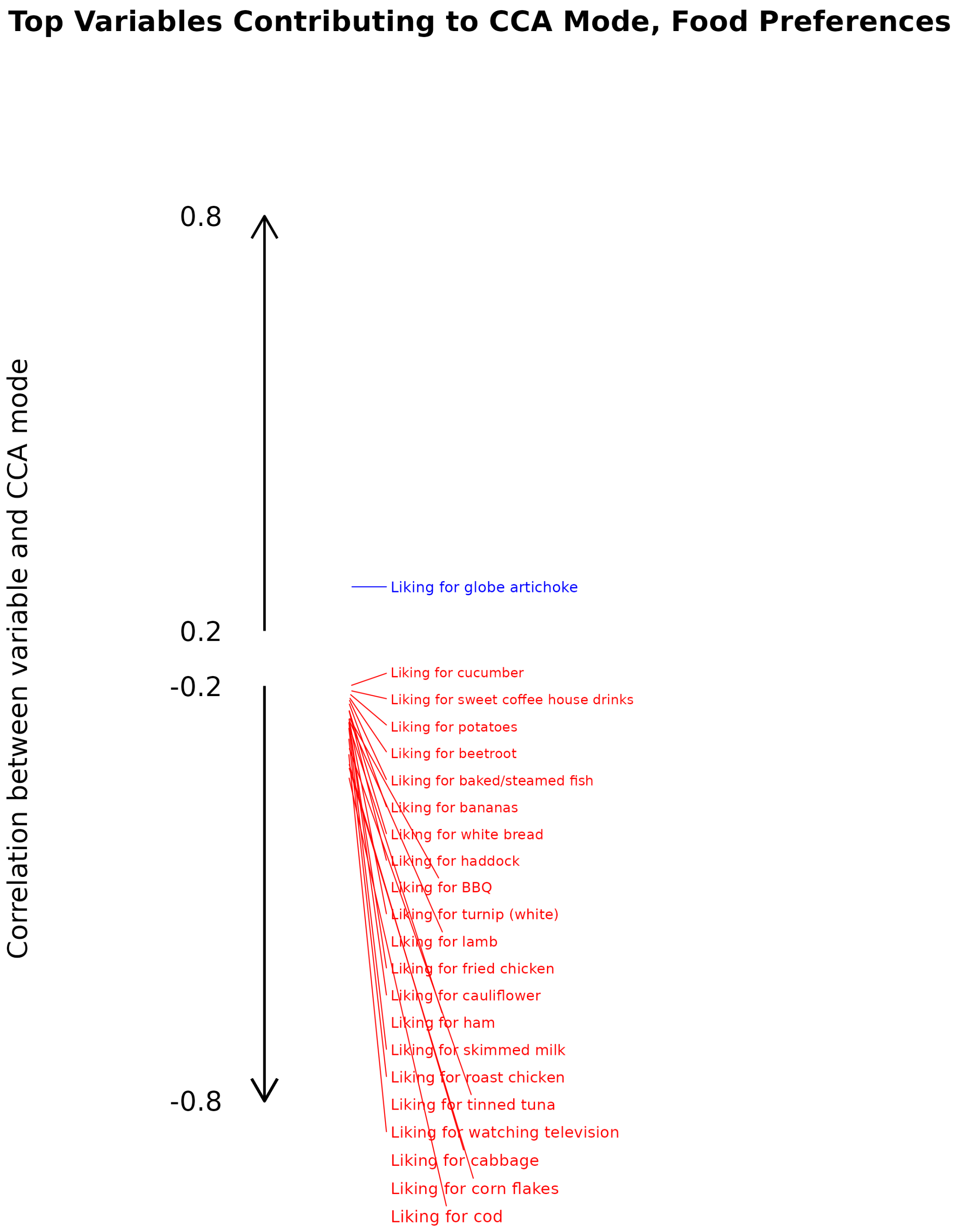


Figure 1: Food preferences: top variables in terms of association with the first canonical mode (IDPs vs Food Preferences). The first canonical correlation is $r = 0.25$.

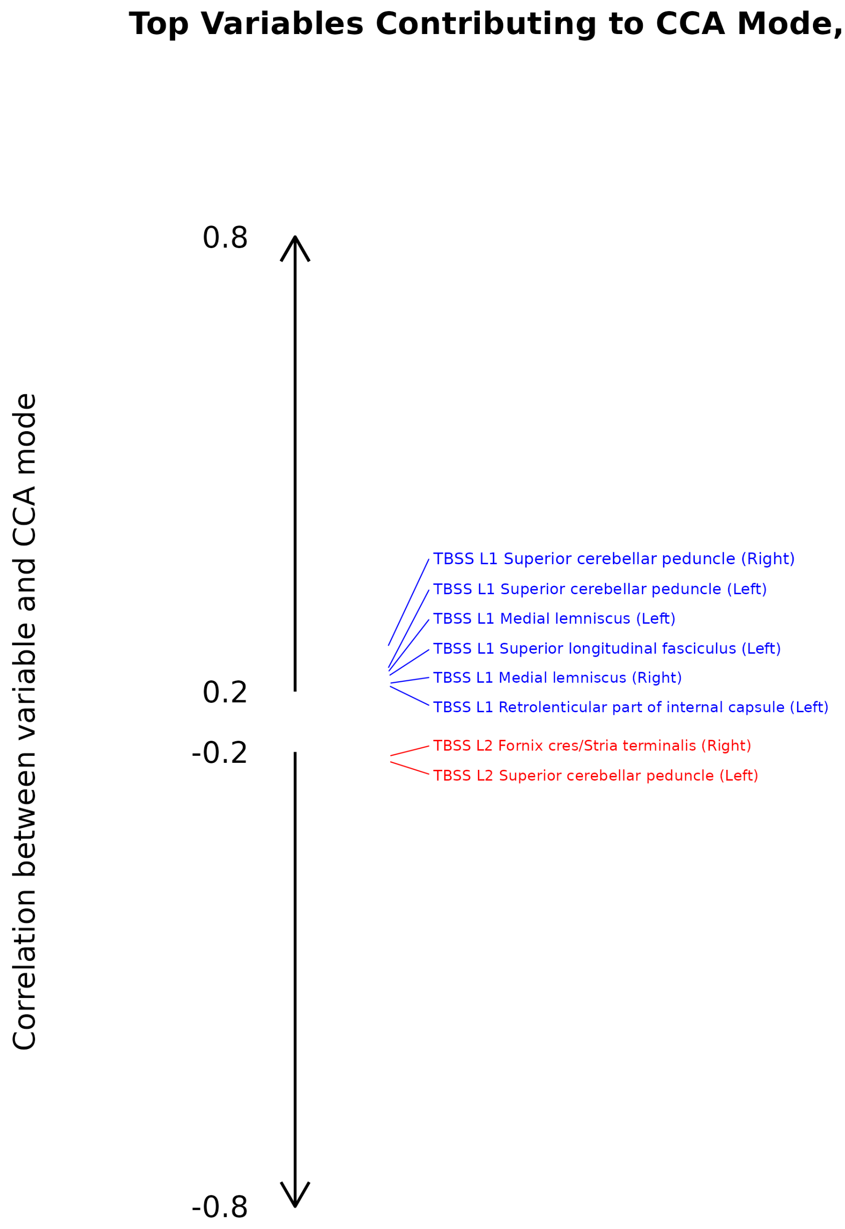


Figure 2: IDPs: top variables in terms of association with the first canonical mode (IDPs vs Food Preferences). The first canonical correlation is $r = 0.25$.

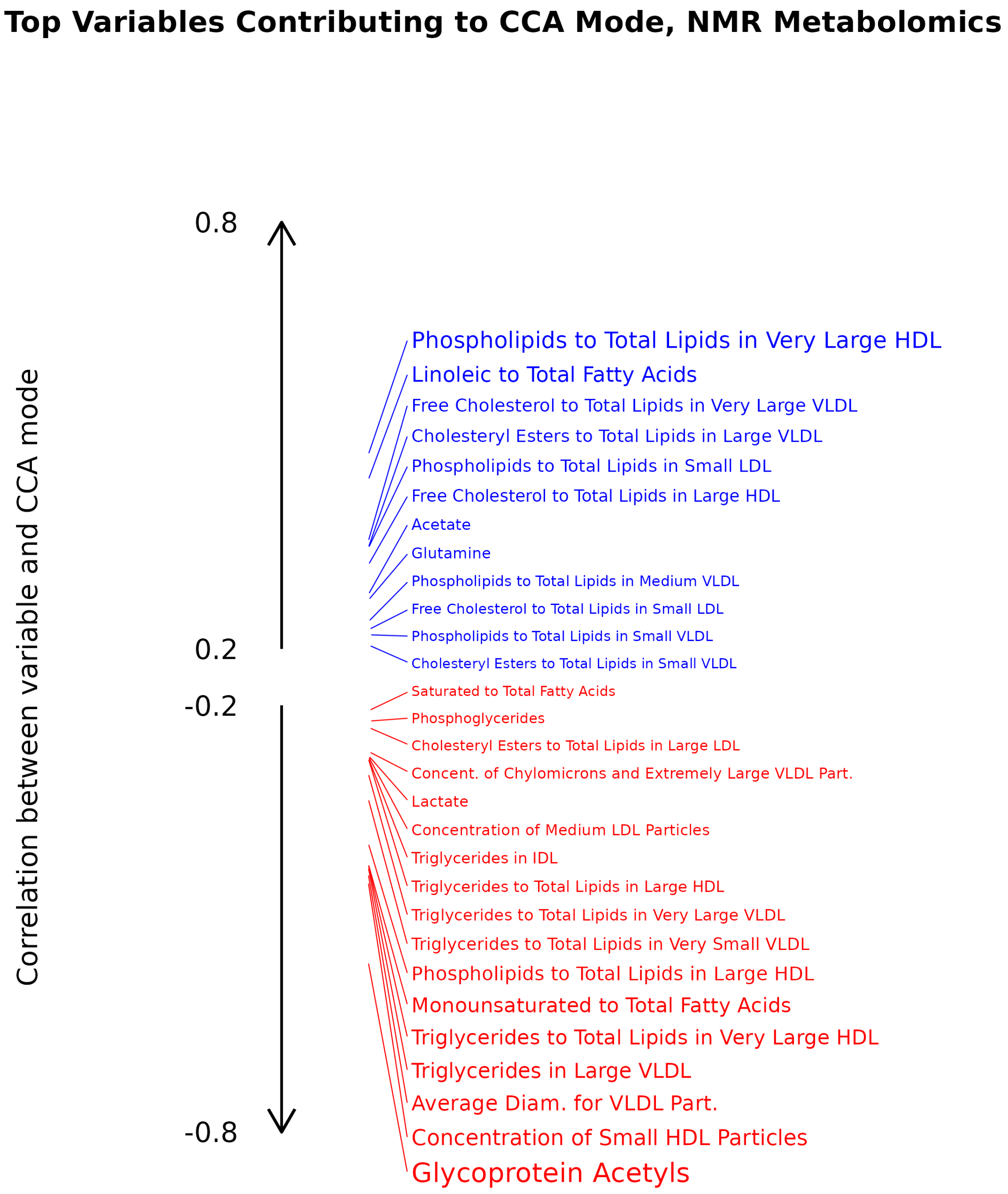


Figure 3: NMR Metabolomics: top variables in terms of association with the first canonical mode (IDPs vs NMR Metabolomics). The first canonical correlation is $r = 0.29$.

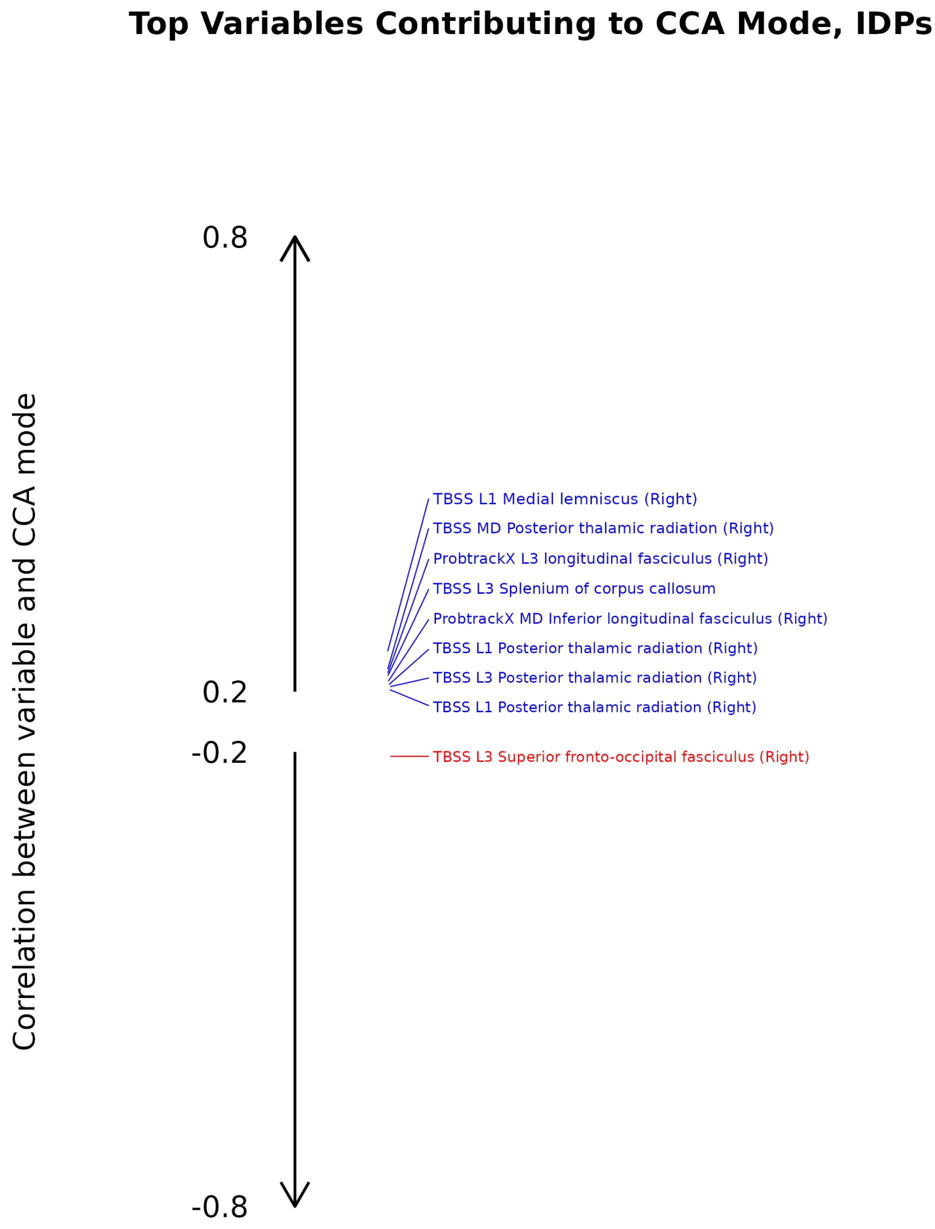


Figure 4: IDPs: top variables in terms of association with the first canonical mode (IDPs vs NMR Metabolomics). The first canonical correlation is $r = 0.29$.

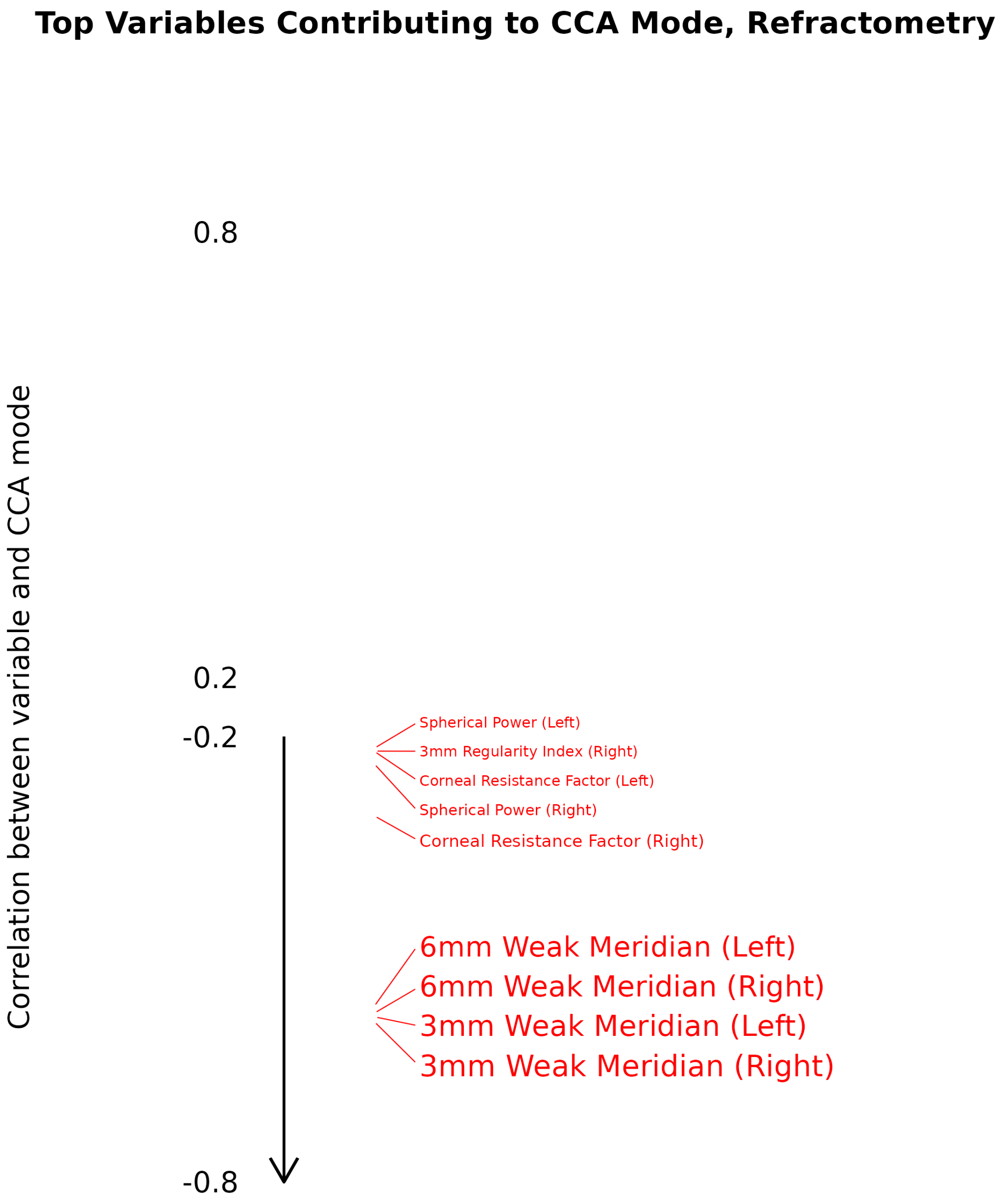


Figure 5: Refractometry: top variables in terms of association with the first canonical mode (IDPs vs Refractometry). The first canonical correlation is $r = 0.28$.

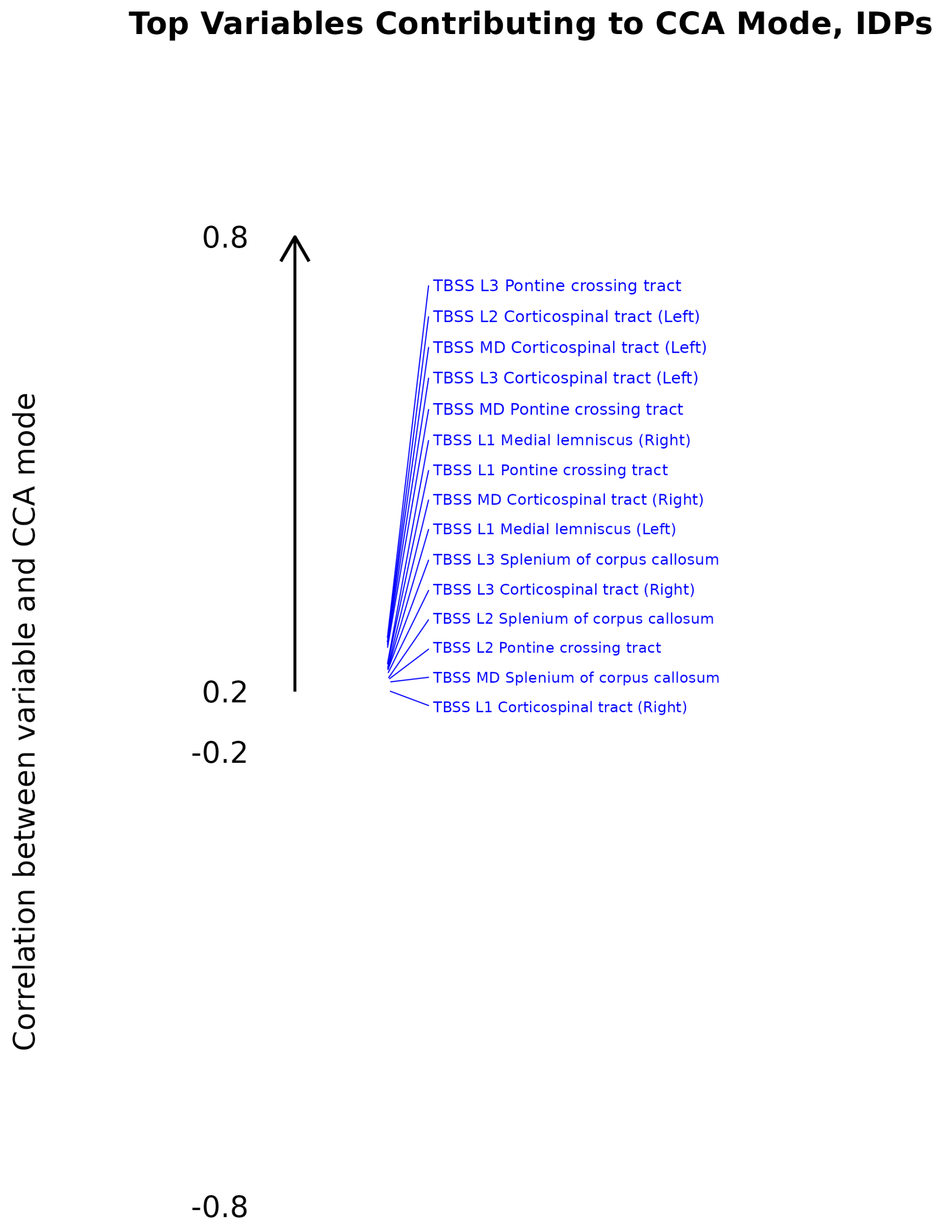


Figure 6: IDPs: top variables in terms of association with the first canonical mode (IDPs vs Refractometry). The first canonical correlation is $r = 0.28$.

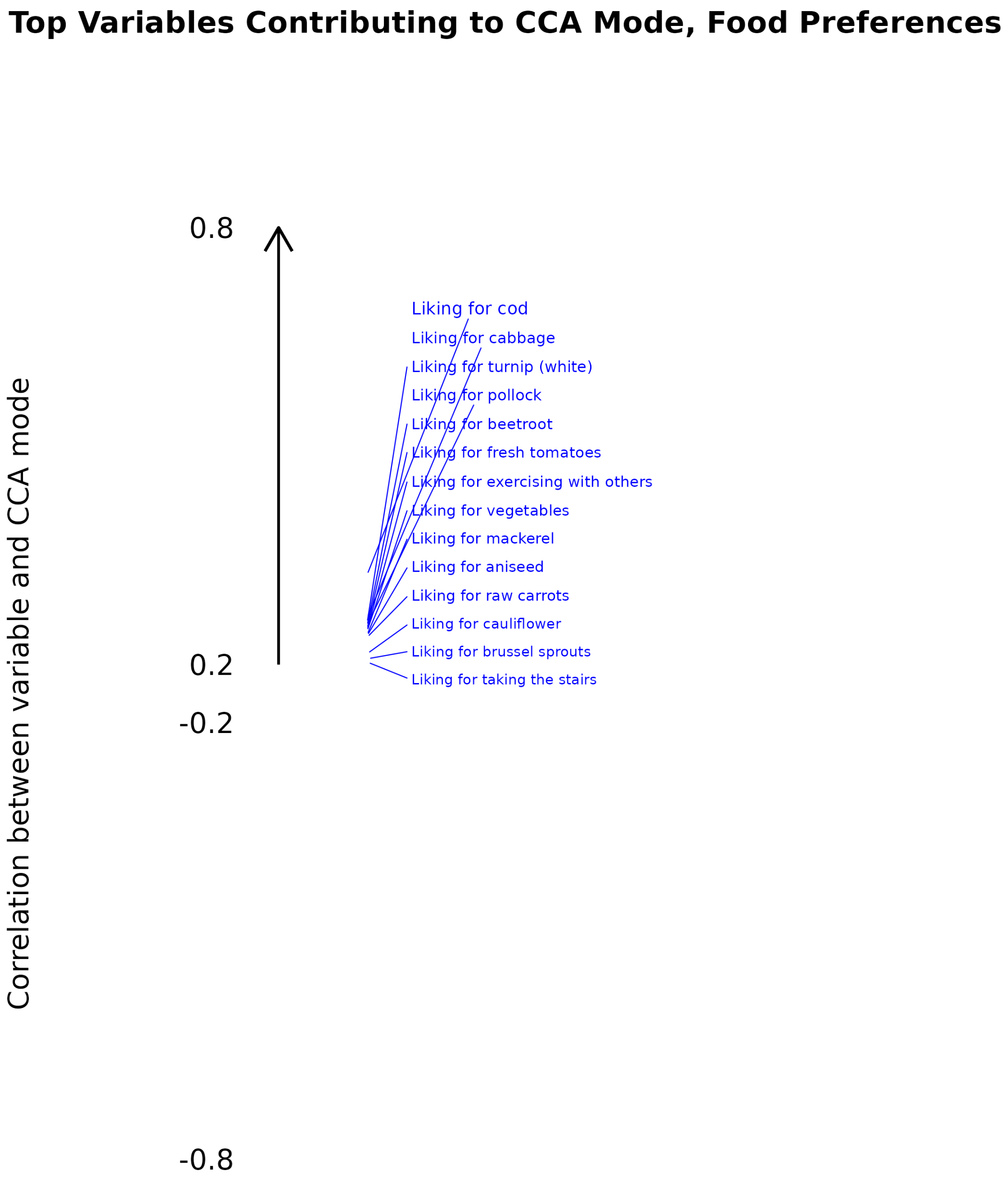


Figure 7: Food Preferences: top variables in terms of association with the first canonical mode (Food Preferences vs Refractometry). The first canonical correlation is $r = 0.23$.

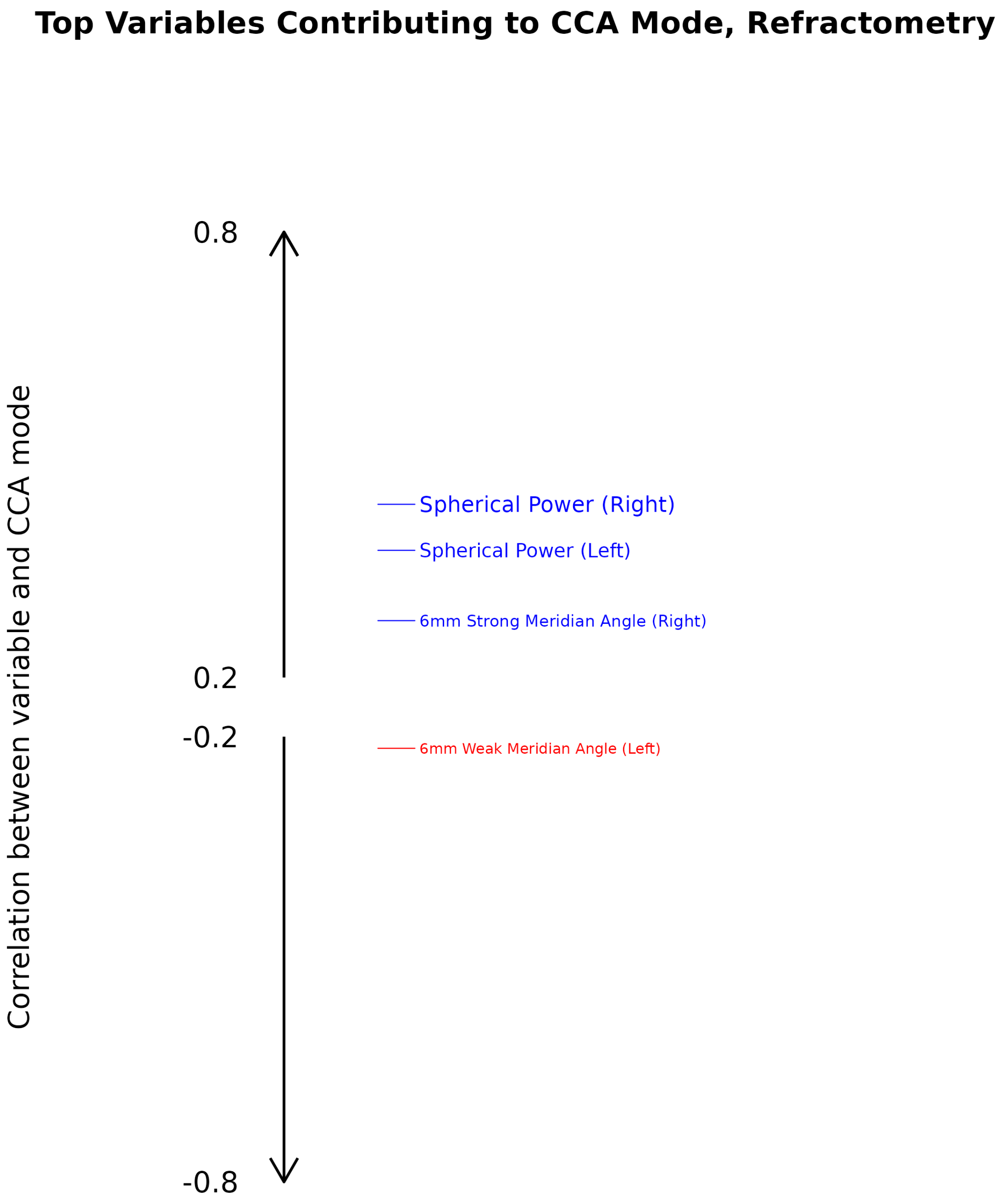


Figure 8: Refractometry: top variables in terms of association with the first canonical mode (Food Preferences vs Refractometry). The first canonical correlation is $r = 0.23$.

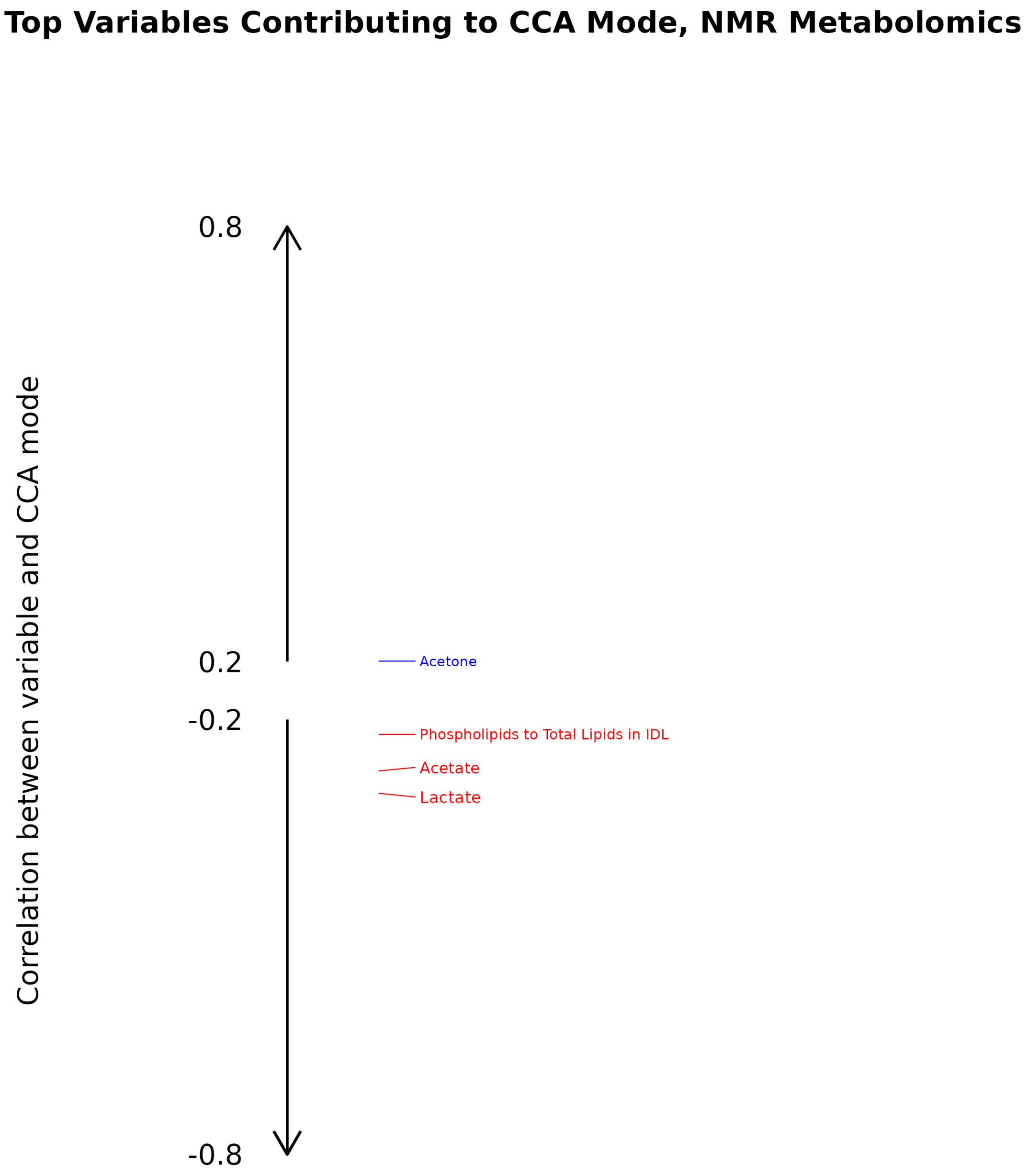


Figure 9: NMR Metabolomics: top variables in terms of association with the first canonical mode (NMR Metabolomics vs Refractometry). The first canonical correlation is $r = 0.42$.

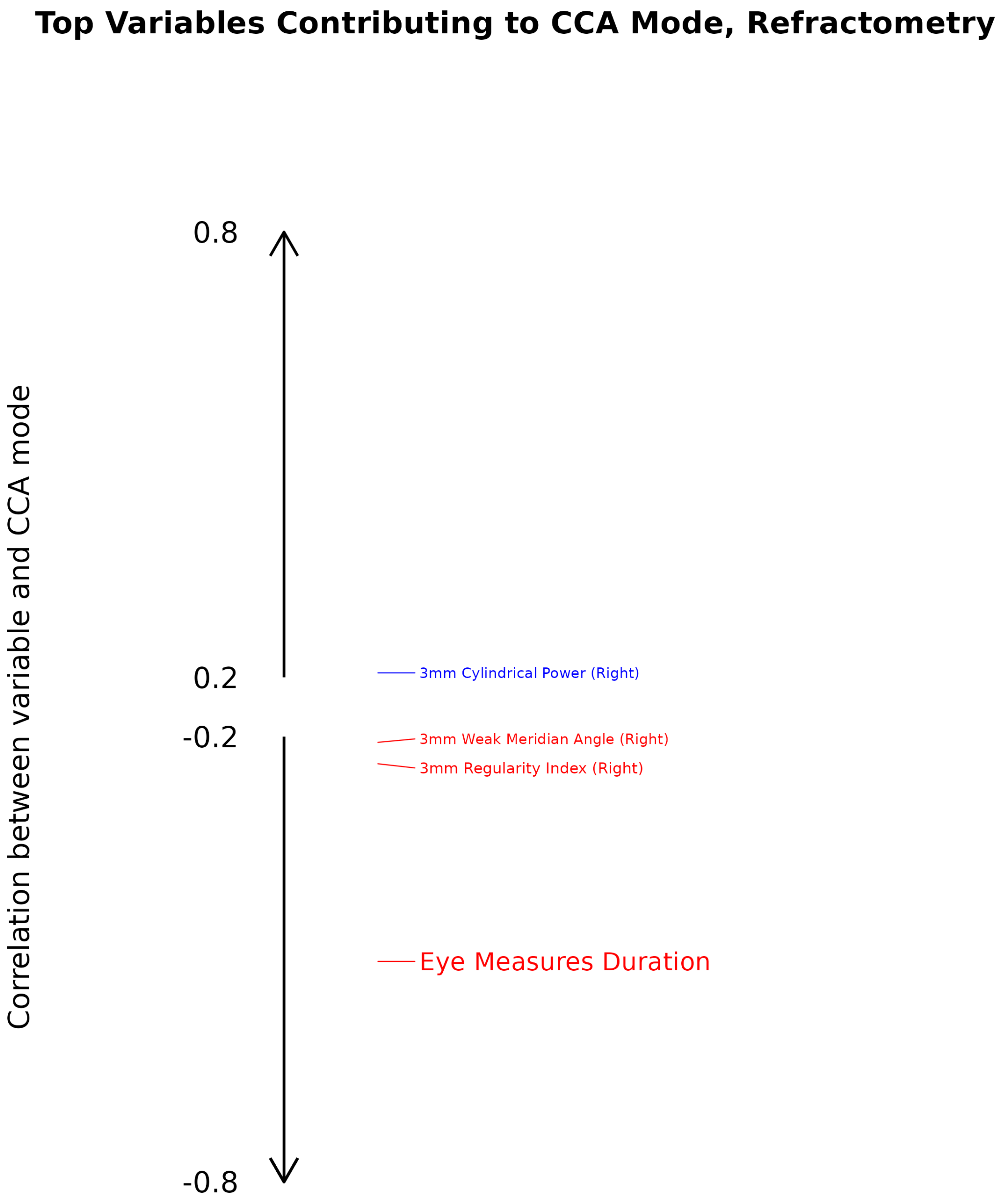


Figure 10: Refractometry: top variables in terms of association with the first canonical mode (NMR Metabolomics vs Refractometry). The first canonical correlation is $r = 0.42$.
